# Untangling age and menopausal status reveals no effect of menopause on white matter hyperintensity volume

**DOI:** 10.1101/2024.10.28.24316270

**Authors:** Denise Wezel, Olivier Parent, Manuela Costantino, Lina Sifi, Grace Pigeau, Nicole J. Gervais, Ann McQuarrie, Josefina Maranzano, Gabriel Allan Devenyi, Mahsa Dadar, M. Mallar Chakravarty

**Author notes:** these authors contributed equally (Denise Wezel, Olivier Parent). **Correspondence to:** M. Mallar Chakravarty, **Address:** 6875 Boulevard LaSalle, Verdun, Montreal, Québec, Canada, H4H 1R3, **Email:**.

## Abstract

**Background and objectives:** White matter hyperintensities (WMHs) are radiological abnormalities indicative of cerebrovascular dysfunction associated with increased risk for cognitive decline and increase in prevalence in older age. However, there are known sex-differences as older females harbour higher WMH burden than males. Some have hypothesized that the increase in this dementia-related risk factor is related to the menopausal transition.

**Methods:** To untangle the effects of age and menopause, we leveraged a large sample from the UK Biobank (n = 10,519) to investigate differences in WMH volumes across the menopausal transition using a strict age-matching procedure.

**Results:** Surprisingly, we find increased WMH volumes in premenopausal women compared to postmenopausal women when simply correcting for age with linear models, but we find no effect in the age-matched sample. Menopause-related characteristics, such as age at menopause or hormone replacement therapy, did not replicate previous literature reporting an association with WMH volumes. Cardiovascular lifestyle variables, such as smoking and blood pressure, were significant predictors of WMH volume in the full sample without age-matching. These effects varied by menopausal status only for days of moderate activity.

**Discussion:** In sum, our findings in a well-powered study suggest that previous reports of menopause-related differences in WMH burden are potentially confounded by age. We further show that the effect of positive lifestyle factors on brain health, as indexed with WMH burden, generally does not change after menopause. Factors other than the menopausal transition are likely at play in explaining the difference in WMH burden between males and females in later life.

## 1. Introduction

White matter hyperintensities (WMHs) are radiological abnormalities appearing on T2-weighted MRI [1] and are recognized markers of small vessel disease and cerebrovascular dysfunction [2]. WMH burden is strongly predicted by age [3] and is associated with maladaptive ageing [4], increased risk for neurovascular pathologies such as stroke [1], cognitive decline and dementia [5], decreased gait speed [6], and overall mortality [1]. Taken together, WMH burden represents a critical neuropathological marker across the neurodegenerative spectrum.

Previous reports show evidence for sex differences in WMH burden. Older females are disproportionately affected by WMHs compared to males of the same age [7–9]. However, these sex differences are not observed in younger samples [9,10], suggesting that they likely arise later in life. The neuroprotective effects of circulating estrogen [11,12] may help explain these consistent observations. Thus, it has been proposed that the increase in WMH burden may be influenced by the menopausal transition [13].

Natural menopause typically occurs between the ages of 45 and 55, and the decrease in ovarian hormones influences various physiological systems, including brain structure and function [14]. This hormonal decrease occurs abruptly in surgically induced menopause, where oophorectomy occurs before the onset of natural menopause transition [15]. The menopausal transition is associated with an increase in cardiovascular risk factors, such as hypertension and diabetes [16,17], that double as risk factors for WMHs [18–20].

Longer periods of ovarian hormone cessation due to a younger age at menopause [21,22] and hot flashes during the menopause transition [23,24], are associated with WMHs. Furthermore, postmenopausal females are reported to have a higher WMH burden than premenopausal females [8,25], and WMHs are reported to increase in recently menopausal females [26]. Consistent with the neuroprotective effects of estrogen, surgical menopause has been associated with increased risk of cognitive impairment and dementia [27] and neurodegeneration [28,29]. However, results are mixed. Previous findings from our group revealed no menopause-related changes in brain volume following a strict age-matching procedure in a large sample [30]. Zeydan et al. (2019) report no difference in WMH burden in surgical compared to natural menopause. Studies of the impact of hormone replacement therapy (HRT) during the menopause transition, often considered to be neuroprotective, remain inconclusive: some studies report mixed conclusions [31,32], while others report no neuroprotective impact of HRT [21,33].

Previous studies examining the relationship between menopause and brain health can be generally categorized into two types: 1) small-sample studies that recruit age-matched controls (usually males) (e.g. [28,34]) or that do not age-match at all [22,26]. 2) larger cohort studies that do not age-match [8,25]. Without age-matching, the difference in average age between the included premenopausal and postmenopausal females can be as much as 14 years in such studies. In some studies, mean age per group is even left unreported. In both designs, disentangling the true neurobiological impact of the menopause transition independent of sex-specific age-related neurodegeneration is challenging. Small sample studies may be underpowered since effect sizes for brain MRI measures are usually small [35]. On the other hand, large population-based studies may find it difficult to recruit age-matched controls.

Here we include a large sample from the UK BioBank and perform the age-matching procedure previously championed by our group [30] to disentangle the effects of age and both surgical and natural menopause on WMHV. We also perform additional analyses to determine if we could replicate findings in the study by Lohner and colleagues (2022), who recently reported significant effects of menopausal status on WMHV in the large population-based Rhineland Study. We further analyze whether WMH burden is influenced by age at self-reported menopause and HRT use and we examine the interplay between lifestyle factors and WMH volumes before and after the menopausal transition. Our findings suggest that reported menopausal transition-related differences in WMH burden are potentially confounded by age and that other factors are therefore likely at play in explaining the difference in WMH burden between males and females in later life. Furthermore, our findings suggest that the effect of lifestyle factors on WMHV generally does not change after menopause.

## 2. Methods

### Sample

The present study uses cross-sectional data obtained from the UK Biobank [36] under application number 45,551. The UK Biobank is a representative population-based study with 39,676 participants from the United Kingdom who underwent brain MRI scans. The full sample included in the present study consists of 10,519 females (self-reported) between 45 and 81 years old with brain MRI data, and is consistent with the sample used in our previous work in Costantino et al. (2023). We assigned participants into three groups: premenopausal females (PRE), females who underwent natural menopause (POST), and females who underwent surgical menopause (bilateral oophorectomy before onset of natural menopause, with or without a hysterectomy; SURG) based on their self report data. Time since menopause was calculated by subtracting age at menopause from age at time of scanning.

Lifetime use of hormone replacement therapy (HRT) was encoded as a binary variable (yes/no). Participants were excluded from the study if they were uncertain of their menopausal status, if they underwent a hysterectomy without a bilateral oophorectomy, if data was missing regarding menopausal status or type, age at menopause, use of hormone therapy, income, or if they reported a diagnosis of multiple sclerosis (MS). In each analysis, different subsamples of the groups were included based on data availability and age matching. Table 1 shows an overview of descriptive statistics for each subgroup. Supplementary table 1 clarifies which dataset is used in which analysis.

**Table 1.**
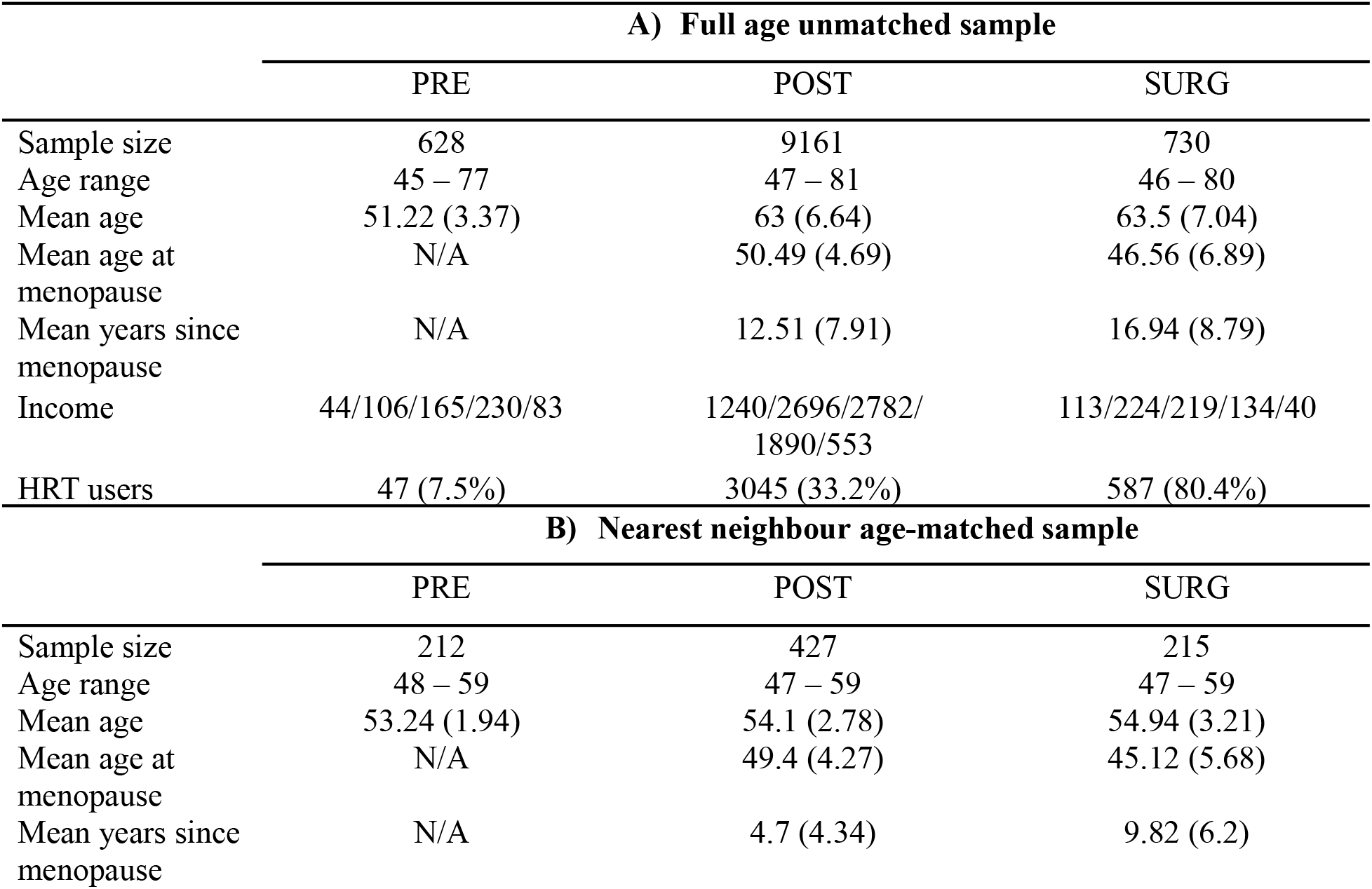
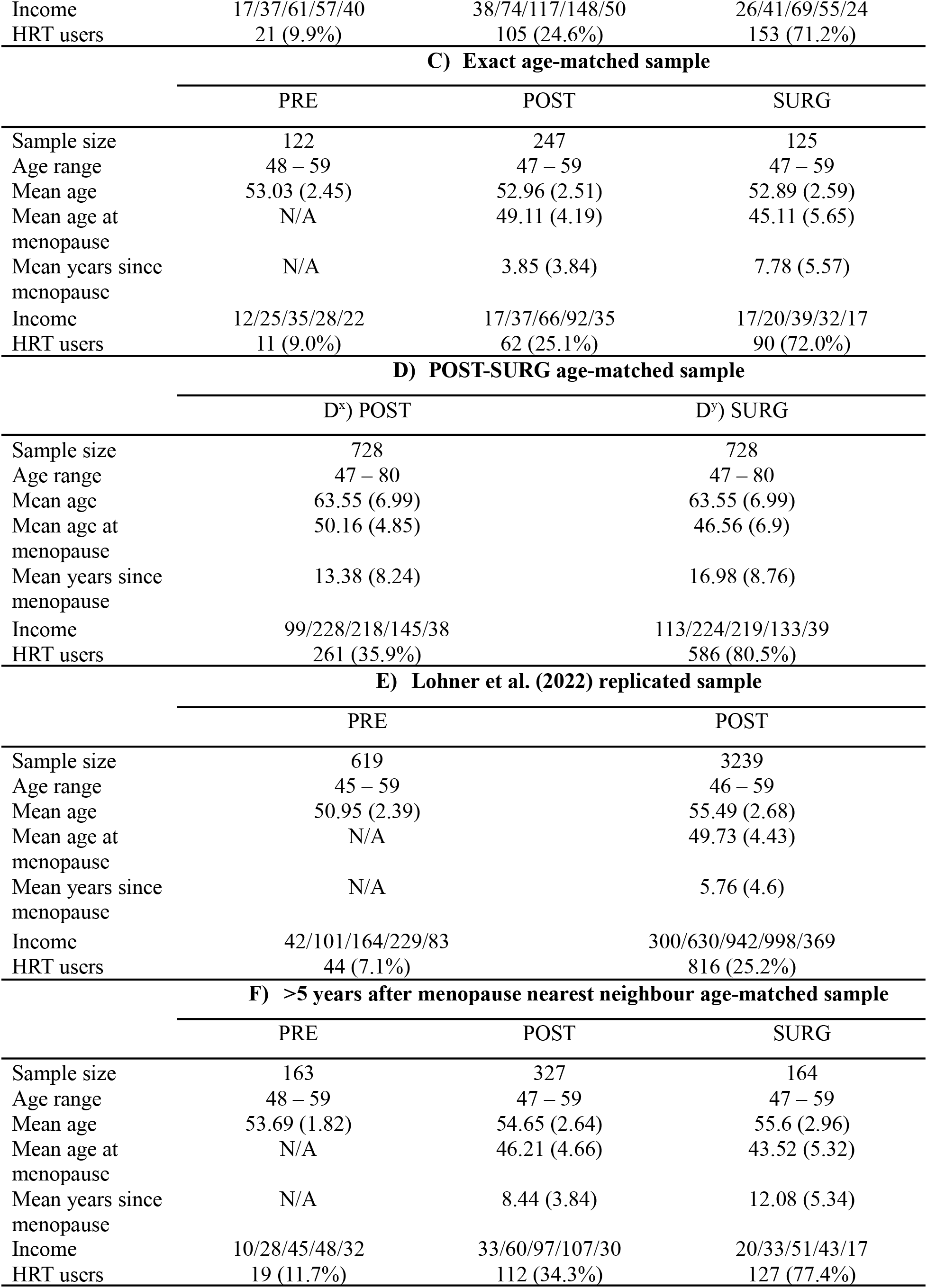
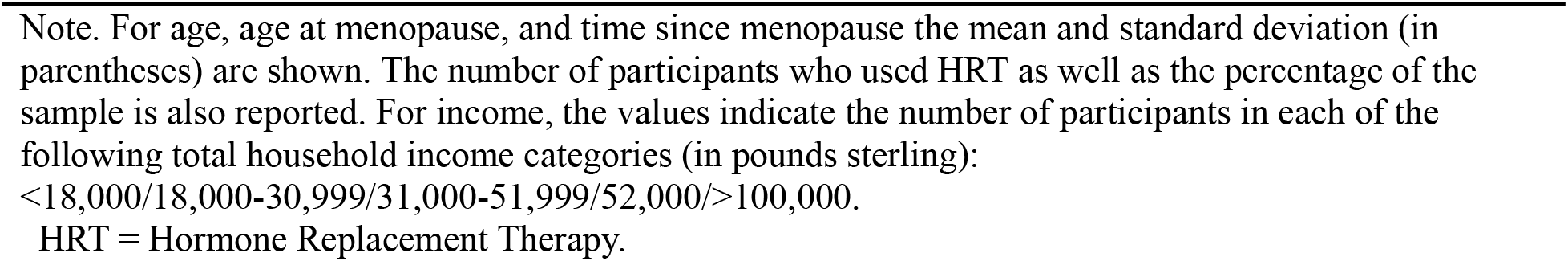
Demographics of participants per group.

### MRI data

All structural MRI data was obtained from the UK Biobank, which were collected from three centers in the United Kingdom using a standardized protocol (Siemens Skyra 3T, with a 32-channel RF receive head coil [37]; see Supplementary Methods 1). T1-weighted (T1w) and fluid-attenuated inversion recovery (FLAIR) structural images were used. While WMH volumes derived by the UK Biobank group are available, we performed multiple processing steps to ensure the highest possible WMH segmentation accuracy (see Supplementary Methods 1). To increase the accuracy of between-subject spatial alignment in the white matter, both T1w and fractional anisotropy maps were used to generate a representative population-based template [38], to which individual participantswere aligned using non-linear registration [39]. Next, we retrained the BraIn SegmentatiON (BISON) algorithm using manually labeled WMHs of 60 UK Biobank participants [40], and then used this trained algorithm to segment WMHs using both T1w and FLAIR as inputs. BISON WMH segmentations were highly correlated with UK Biobank-derived values computed with BIANCA from FSL (*r* = 0.86, *p* < 0.001) and in cases of discrepancy, visual assessment favored BISON segmentations. This procedure thus ensured a high segmentation accuracy of WMHs. An example of WMH segmentation can be found in figure 1A.

**Figure 1.**
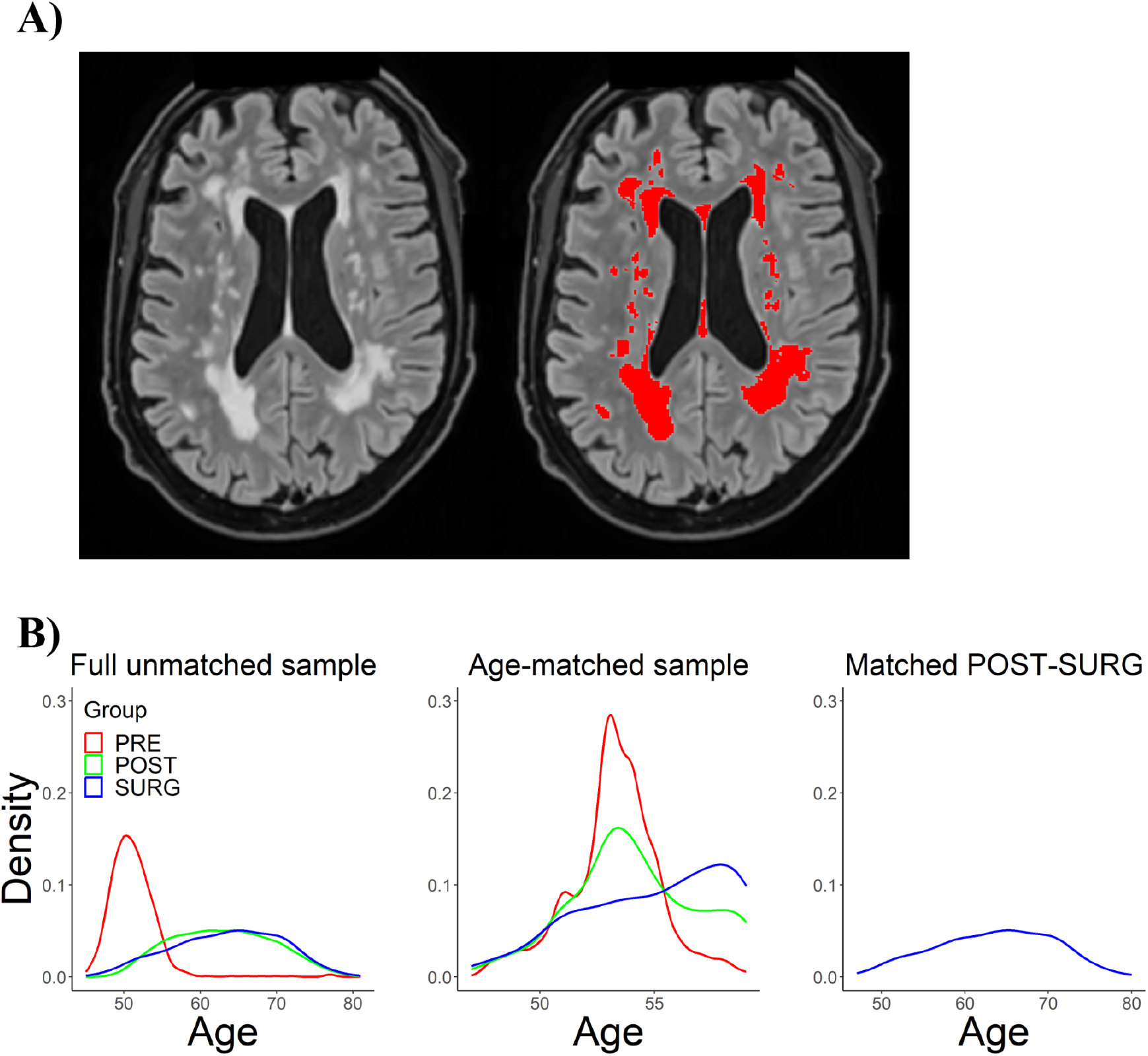
**A)** shows an example of WMH segmentation from the UK Biobank. The left image shows the preprocessed FLAIR image, the right image shows the BISON WMH mask. **B)** shows the age distributions before and after age-matching. The middle panel shows the nearest neighbour age-matched sample.

Manual quality control of raw T1-weighted scans was done following guidelines developed by our group (https://github.com/CoBrALab/documentation/wiki/Motion-Quality-Control-(QC)-Manual; [30,41]. WMHV values were divided by total brain volume (using UK Biobank provided volumes) and log-transformed to address skewness, as suggested by previous studies [42].

### Lifestyle data

Lifestyle risk factors were selected from the available UK Biobank data to be representative measures of known cardiovascular risk factors for WMHs and dementia [18,19,43]. We included a total of 17 variables in the categories of alcohol consumption, blood pressure, diabetes, obesity, physical activity, and smoking. Distributions of variables were visually inspected and, where needed, variables were calculated, binarized, log-transformed, or averaged. When necessary, data were recoded so that higher values indicated overall higher burden. For example in data field 1558 (alcohol intake frequency), the initial data was coded on a 6-point scale where less alcohol intake had the highest value. We recoded this variable so that a lower alcohol intake yielded a value of 1 and the highest value possible was 6. A full overview of all included lifestyle variables, their corresponding UK Biobank Field IDs, and the performed transformations can be found in supplementary table 2. Total household income before tax was used to control for income.

### Statistical analyses

All statistical analyses were performed using R (version 4.1.2). Linear models were used for all statistical tests using z-scored continuous variables to obtain standardized coefficients. Each analysis included age, HRT, and income, and multiple comparisons and were corrected for using false discovery rate (FDR) correction [44,45].

#### Samples

Age-matched groups were created as in Costantino et al. (2023), using the k-nearest neighbors algorithm from the MatchIt package in R [46]. Propensity scores based on the generalized linear model were used to estimate distance between individuals.

The full unmatched sample consists of all individuals that met the inclusion criteria (table 1A). To create the nearest neighbour age-matched sample (table 1B) the PRE group was used as a reference and the SURG group was matched to this group. After this, the POST group was matched to the resulting participants using the same method. Figure 1B shows a comparison of the density plots before and after age-matching. The exact age-matched sample (table 1C) was created using the same method, but an exact match was required instead of the nearest match, making the age-matching more strict to rule out any artifactual results due to the matching technique. For the POST-SURG age-matched sample (table 1D), the POST and SURG groups were matched directly. These same age-matched individuals were included when analyzing POST and SURG groups separately (table 1D^x^ and 1D^y^).

Most analyses were done using the aforementioned three datasets. We performed two additional analyses. First, we attempted to replicate the study by Lohner and colleagues (2022). To create the sample for this analysis we matched their processing steps by normalizing WMHV using white matter volume (provided by UK Biobank) instead of total brain volume, restricting the age range by excluding participants over 59, and not differentiating between natural and surgical menopause (table 1E). Second, as the loss of estrogens could theoretically take time to influence vascular brain health, we created a sample by first excluding all participants who had menopause less than 5 years prior to scanning to keep within the bounds of our age-matched dataset, and then age-matched using the same method as for the nearest neighbour age-matched sample (table 1F).

#### Differences in WMHV across the menopausal transition

First, to investigate whether menopausal groups differ in terms of WMHV, we analysed the nearest neighbour age-matched sample. Additionally, the same model was run in the exact age-matched sample and in the full unmatched sample (adding age as a covariate).

Second, in an attempt to replicate a similar analysis performed in a study by Lohner and colleagues (2022), we matched their analysis scheme by including cardiovascular covariates (hypertension, smoking, and BMI).

Third, we analyzed whether there were differences in WMHV across groups after excluding participants who had menopause less than 5 years prior to scanning.

#### Effect of age at menopause and HRT use on WMHV

We examined the association between age at menopause and WMHV (in the POST and SURG samples separately) to examine potential effects of age at menopause, as well as the interaction between group and age at menopause (in the POST-SURG age-matched sample) to explore whether the effect of age at menopause on WMHV is different in natural and surgical menopause. We tested the relationship between HRT and WMHV (in the POST and SURG samples separately), as well as the interactions of HRT by group (in the POST-SURG age-matched sample). We also examined the interaction of HRT by age at menopause (in the POST and SURG samples separately).

#### Effect of menopausal status on the relationships between lifestyle factors and WMHV

We investigated the effect of lifestyle cardiovascular factors on WMHs across the menopausal transition. To confirm the previously reported effects of lifestyle variables on WMHV [18–20], we assessed these relationships in the full unmatched sample. Further, we explored lifestyle variable by group interactions in the nearest neighbour age-matched sample to assess whether the suggested protective effect of a healthy lifestyle depends on the menopausal status. Additionally, we examined whether usage of blood pressure medication has an effect on WMHV and whether this effect was the same in the different menopausal groups (analysis in the nearest neighbour age-matched sample). A full list of all models can be found in supplementary table 1.

## 3. Results

After manual quality control of the raw MRI images, 4113 participants were excluded (342 (35%) in the PRE group, 3405 (27%) in the POST group, and 366 (33%) in the SURG group). A full description of the samples is displayed in table 1, and an overview of which datasets were used for the different models is displayed in supplementary table 1.

### Differences in WMHV across the menopausal transition

In the full unmatched sample, linear models including age as a covariate showed significant differences between the menopausal groups in terms of WMHV (POST: *β* = −.14, *p* < .001; SURG: *β* = −.15, *p* = .003), revealing that postmenopausal females and females who underwent surgical menopause have lower WMHV than premenopausal females, which is contrary to previous findings [8,25] (figure 2A). However, in the nearest neighbour age-matched sample there were no significant differences between groups (POST: *β* = −.07, *p* = .394; SURG: *β* = −.04, *p* = .678) (figure 2B; figure 2E). Similarly, in the exact age-matched sample, there were also no differences between groups (POST: *β* = −.10, *p* = .367; SURG: *β* = −.11, *p* = .422).

**Figure 2.**
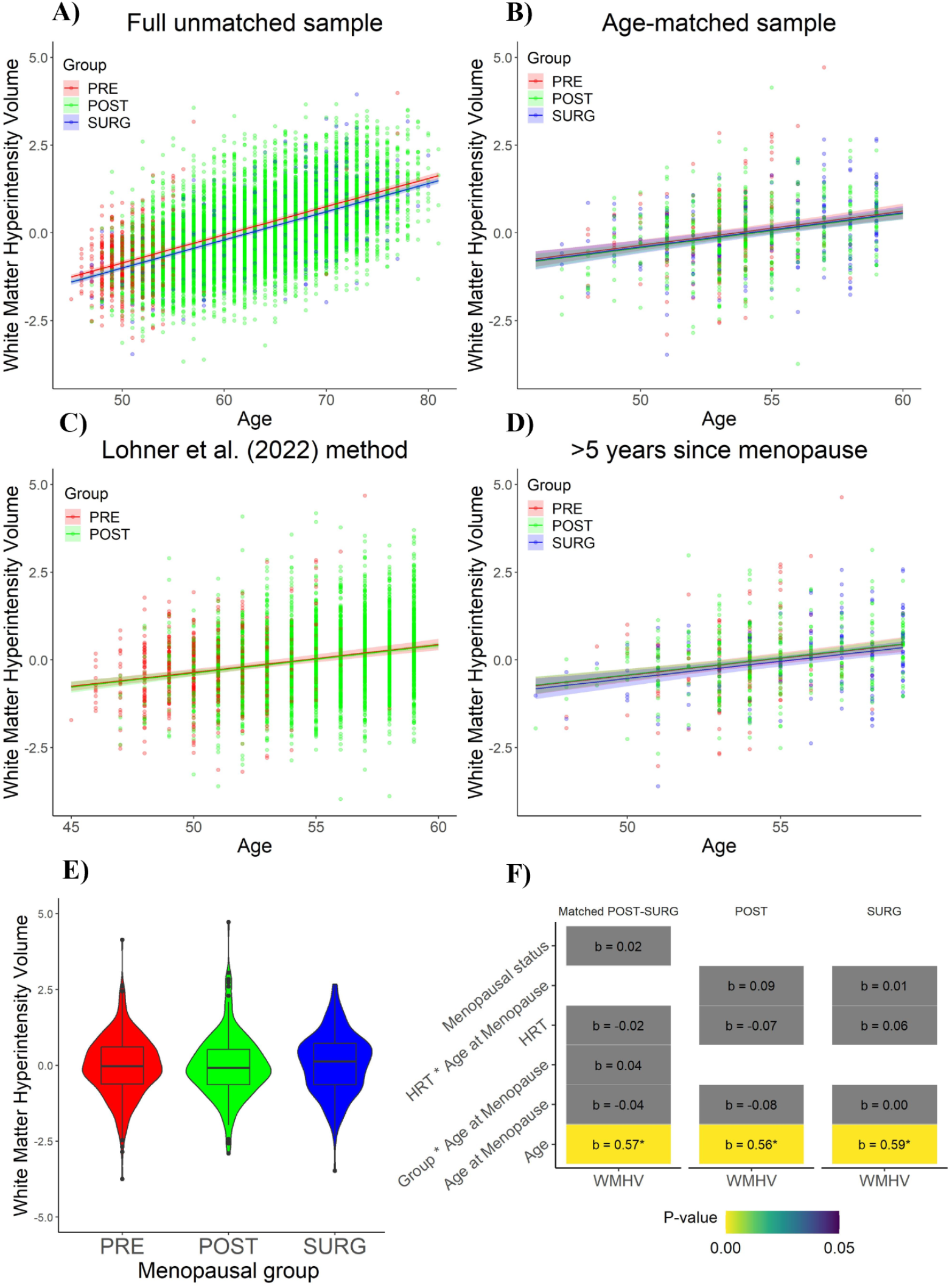
Results of linear models. **A)** - **D)** Line graphs showing the relationship between age and WMHV, separated by group, in the four different models. Panel B shows the nearest neighbour age-matched sample. **E**) Violin plot showing the absence of differences across menopausal groups in white matter hyperintensity volumes in the age-matched sample. **F**) Summary of the results of the linear models that were run. The number in the cell represents the standardized beta, with an asterisk marking that this effect survives FDR-correction. The color of the cells represent the p-value, with gray cells representing a p-value over .05. Income was corrected for in each model.

When using methods similar to Lohner and colleagues (2022), we also found no significant group differences (*β* = −.02, *p* = .714) (figure 2C). Furthermore, the supplementary analysis in which only participants who had menopause at least 5 years prior to scanning were included showed no group differences (POST: *β* = 0.02, *p* = .867; SURG: *β* = −.09, *p* = .426) (figure 2D).

In sum, we did not detect that post-menopausal females have higher WMHV than premenopausal females when accounting for age using four different analysis strategies or when using analytical strategies previously used by other groups.

### Effect of age at menopause and HRT use on WMHV

No associations were found between age at menopause and WMHV within the POST (*β* = −.03, *p* = .304) and SURG (*β* = .01, *p* = .756) group. Furthermore, no group by age at menopause interaction was found (*β* = .04, *p* = .394) (analysis in the POST-SURG age-matched sample) (figure 2F).

No main effect was found for HRT in the nearest neighbour age-matched sample (*β* = .02, *p* = .779), or in the separate POST (*β* = −.07, *p* = .310) and SURG (*β* = .06, *p* = .417) groups (table D^x^ and D^y^) (figure 2F). Furthermore, no HRT by group interaction (*β* = −.06, *p* = .451) or HRT by age at menopause interaction (POST group: *β* = .09, *p* = .156; SURG group: *β* = .01, *p* = .928) were found (analysis in the nearest neighbour age-matched sample) (figure 2F).

In sum, we did not detect that age at menopause or use of HRT influenced WMHV, whether within or differently between menopausal groups.

### Effect of menopausal status on the relationships between lifestyle factors and WMHV

All lifestyle variables tested except those related to physical activity were significant predictors of WMHV in the full unmatched sample (figure 3A). In the nearest neighbour age-matched sample, a group by lifestyle variable interaction was only found for one lifestyle variable (number of days of moderate physical activity) (figure 3B). Furthermore, we found no group by blood pressure medication interaction (POST: *β* = −.11, *p* = .771; SURG: *β* = −.25, *p* = .412), while controlling for diastolic blood pressure, systolic blood pressure, and hypertension.

**Figure 3.**
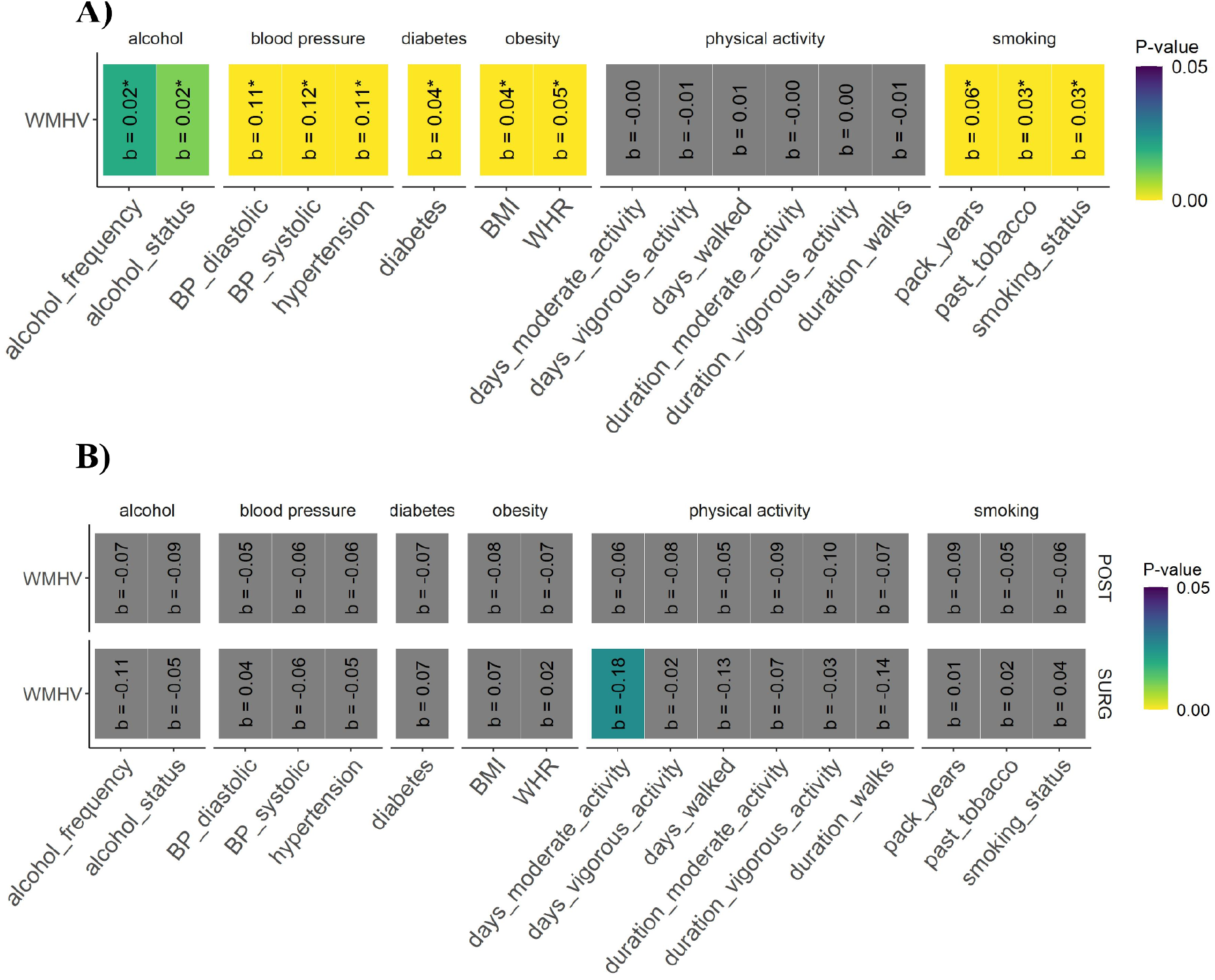
Effects of menopausal status on the relationships between lifestyle factors and WMHV. The number in each cell represents the standardized beta, with an asterisk marking that this effect survives at FDR<0.05. The color of the cells represent the p-value, with gray cells representing a p-value over .05. Each model was corrected for SES, age, and HRT. **A)** shows the results of the linear models that were run in the full unmatched sample. **B)** shows the results of the linear models with the group by lifestyle variable interaction terms that were run in the nearest neighbour age-matched sample.

In sum, we did not detect that the positive influence of a healthy lifestyle or blood pressure medication on WMHV differed across the menopausal transition.

## 4. Discussion

In the present study, we investigated the relationships between WMHV and menopausal status and type, age at menopause, HRT, and lifestyle variables in a large, well-powered sample obtained from the UK Biobank. Overall, the findings suggest that there are no WMHV differences between premenopausal females, postmenopausal females, and females who underwent surgical menopause in an age-matched sample. Interestingly, without age-matching but only controlling for age, the results did show group differences, but in the opposite direction previously reported in the literature (post-menopausal women had *lower* WMHV than pre-menopausal women) [8,25]. A potential explanation is that controlling for age is not enough to fully untangle the effects of age and menopausal status. Since the age distributions of the pre- and post-menopausal groups are largely non-overlapping in the full unmatched sample, the age covariate would encode and extrapolate the effect for both groups, which could lead to inaccurate correction. For this reason, we consider that our age-matching procedure more accurately removed the effect of age.

We also attempted to replicate one of the studies that has reported that postmenopausal females have higher WMH burden than premenopausal females [8], but did not find an effect, even without age-matching. Intriguingly, the current study and the one by Lohner and colleagues (2022) have similar samples (UK Biobank and the Rhineland Study, respectively), both being community-based studies in Europe and having similar sample sizes of pre- and post-menopausal women (in our age-matched sample). While our opposite findings could simply be caused by random sample variability, it is possible that our more stringent age-matching procedure better isolated the effect of menopausal status (or the lack thereof). Furthermore, in individuals who had menopause at least 5 years prior to scanning, no group differences were found, which shows that even after time has passed post-menopause no differences emerge. The choices that were made to isolate the impact of menopause were in part necessary because the UK Biobank was not solely developed to study the effects of menopause on other biological variables. Future prospective studies that are adequately designed and powered to specifically isolate the impact of all stages of the menopause transition on biological phenotypes are urgently needed.

Contrary to previous studies, no effect of age at menopause was found in any group. However, these previous studies had small samples [22], and used categorical instead of continuous measure for age at menopause [21,22]. Furthermore, we found no effect of HRT on WMHV, which is consistent with other literature that has reported mixed or negative findings on the protective effects of HRT on brain health [21,32,33]. However, due to the absence of data regarding type, dosage and duration of HRT in the UK Biobank, we cannot exclude a more subtle, dose-dependent effect of HRT use on WMHV [31,47] and further research is needed to gain a more comprehensive understanding of its potential effects.

Our findings support the relationship between cardiovascular risk factors and WMHV. Specifically, the risk factors linked to WMHV in the present study were in the categories of alcohol consumption, blood pressure, diabetes, obesity, and smoking. Interestingly, contrary to previous reports [48], no effects were observed for physical activity, implying a potential absence of a direct relationship between physical activity and WMHV. Notably, [43] reported that physical inactivity is a risk factor for dementia. However, Habes and colleagues (2016) also reported no significant interactions between physical activity and WMHV. This suggests that while physical activity may be a risk factor for general brain aging and dementia, there may not be an association with WMHV specifically. However, another possible explanation could be that the physical activity data in the UK Biobank reflects the current level of activity, which may not accurately represent the physical activity patterns throughout life. A measure of activity throughout the lifespan may be more suitable for drawing conclusions. The effect of lifestyle variables and blood pressure medication on WMHV were not statistically different before and after the menopausal transition. We therefore provide evidence that the protective effects of a healthy lifestyle on brain health, using WMHV as a proxy, do not change after the menopausal transition.

The present study should be considered in light of its limitations. Firstly, the UK Biobank contains no data on perimenopause, which may have led to mislabeling of individuals in the premenopausal group. Given that perimenopause typically lasts around 4 years [49] and is associated with fluctuations of sex hormone levels [50], the inclusion of perimenopausal individuals within the premenopausal group could have biased the groupings. No conclusions can be made on perimenopausal females based on these findings. Furthermore, data on menopause and age at menopause relied on self-report, raising uncertainties about the reliability of this data. For instance, older individuals who experienced menopause decades ago may not be able to accurately recall their precise age at menopause. However, most of these individuals will not have been included in our younger, age-matched sample. Lastly, the UK Biobank sample is not gender diverse (i.e. no transgender or intersex individuals are explicitly included and no self-identification questionnaires regarding gender were used) and consists predominantly of self-reported as ‘White British people’, thus our findings may not be generalizable to a broad, diverse population in terms of gender, culture, nationality, or ethnicity.

Despite these limitations, the present study is one of most extensive and well-powered in its field. We took great care in the quality of the MRI data by excluding participants that had motion artifacts and building a custom processing pipeline to maximize WMH segmentation accuracy. A large sample was included and a robust age-matching approach allowed us to accurately assess the trends of WMHV across the menopausal transition. We demonstrated that by isolating the effect of menopausal transition from aging, there appears to be no impact of the menopausal transition on WMH burden. Consequently, these findings underline the importance of employing age-matching methods in future studies rather than relying only on statistically controlling for age, particularly when groups have largely non-overlapping age distributions. Moreover, we showed that the protective effects of lifestyle factors on WMHV do not change after menopause, which emphasizes the importance of maintaining a healthy lifestyle throughout life to preserve brain health. Contrary to conclusions of previous studies, our results suggest that factors other than the menopausal transition are likely at play in explaining the difference in WMH burden between males and females in later life. This insight opens avenues for further investigation into the underlying mechanisms that contribute to these sex differences.

In summary, our study contributes important knowledge to the field and to female-specific health factors in aging, highlighting the interplay between the menopausal transition, brain health, and cardiovascular risk factors. Research has historically overlooked female health, and this study adds to the body of literature that aims to fill this critical gap. Furthermore, our study underscores the importance of methodological considerations in future research and emphasizes the importance of a healthy lifestyle throughout life for maintaining a healthy brain.

## Data Availability

Requests for access to the data used for this study will be considered by the corresponding author.

