## Supplementary materials for "Untangling age and menopausal status reveals no effect of menopause on white matter hyperintensity volume"

### Supplementary methods 1 - Image processing

First, both the raw T1w and FLAIR images were denoised [1], field-inhomogeneity corrected [2], and intensity normalized between 0 and 100. A brain mask was computed with the BEaST algorithm [3]. The FLAIR images were rigidly registered to the T1w images with the Advanced Normalization Tools (ANTs) pipeline [4]. Second, to ensure high registration accuracy to a common template space, we generated a custom UK Biobank template with the T1w images of 100 males and 100 females with representative age distributions [5]. Third, to enhance the registration accuracy in the white matter tracts, which has low contrast on T1w images, we leveraged the fractional anisotropy (FA) maps derived from diffusion-weighted imaging [6], which were further denoised, super-sampled to 1 mm isotropic in resolution [7], and rigidly registered to the T1w image [4]. We then performed multispectral non-linear registration, using processed T1w and FA images as inputs, to register to the common template. Fourth, we manually labeled the WMHs of 60 UK Biobank participants, which we then used to retrain the validated Brain Tissue Segmentation (BISON) pipeline [8]. Applied to the whole cohort, this segmented the brain into 9 tissue types including WMHs using both preprocessed T1w and FLAIR images as inputs (https://github.com/VANDAlab/BISON-WMH).

**Supplementary table 1**
*Overview of all models and datasets used*

| **Differences in WMHV across the menopausal transition** | |
|---|---|
| Model | Sample(s) |
| WMHV ~ group + age + income + HRT | A, B, C, F |
| WMHV ~ group + age + income + hypertension + smoking status + past tobacco + pack years + BMI | E |
| **Effect of age at menopause and HRT use on WMHV** | |
| Model | Sample(s) |
| WMHV ~ age at menopause + age + income + HRT | $D^x$, $D^y$ |
| WMHV ~ group * age at menopause + age + income + HRT | D |
| WMHV ~ HRT + group + age + income + age at menopause | D, |
| WMHV ~ HRT + age + income + age at menopause | $D^x$, $D^y$ |
| WMHV ~ HRT * group + age + income | D |
| WMHV ~ HRT * age at menopause + age + income | $D^x$, $D^y$ |
| **Effect of menopausal status on the relationships between lifestyle factors and WMHV** | |
| Model | Sample(s) |
| WMHV ~ lifestyle variable + age + income + HRT | A |
| WMHV ~ lifestyle variable * group + age + income + HRT | B |
| WMHV ~ group * BP medication + diastolic blood pressure + systolic blood pressure + age + income | B |

Note. Table shows all models that were run, in order of mention in the results section. Models were run using different datasets, which is specified by the letter behind the model (A: full unmatched sample, B: nearest neighbour age-matched sample, C: exact age-matched sample, D: POST-SURG age-matched, $D^x$: POST, $D^y$: SURG, E: Lohner et al. (2022) replication sample, F: >5 years after menopause age-matched. All samples are described in table 1.

**Supplementary table 2**

*Risk factor variables.*

| Category | Name | Field ID | Variable name | Transformation |
|---|---|---|---|---|
| Alcohol | Alcohol drinker status | 20117 | alcohol_status | Binarized: never and previous combined into no |
| | Alcohol intake frequency | 1558 | alcohol_frequency | Reverse coding |
| Blood pressure | Regularly takes blood pressure medication | 6153 | BP_medication | Binarized to yes and no for blood pressure medication |
| | Diastolic blood pressure | - | BP_diastolic | Calculated by averaging data fields 94 and 4079 |
| | Systolic blood pressure | - | BP_systolic | Calculated by averaging data fields 93 and 4080 |
| Diabetes | Diabetes diagnosed by doctor | 2443 | diabetes | - |
| Obesity | Body mass index (BMI) | 23104 | BMI | - |
| | Waist to hip ratio (WHR) | - | WHR | Calculated using waist circumference (data field 48) and hip circumference (data field 49) |
| Physical activity | Number of days/week walked 10+ minutes | 864 | days_walked | - |
| | Duration of walks | 874 | duration_walks | Collapsed at 3.5 SD above mean, log-transformed |
| | Number of days/week of moderate physical activity 10+ minutes | 884 | days_moderate_activity | - |

| | | | | |
|---|---|---|---|---|
| | Duration of moderate activity | 894 | duration_moderate_activity | Collapsed at 3.5 SD above mean, log-transformed |
| | Number of days/week of vigorous physical activity 10+ minutes | 904 | days_vigorous_activity | - |
| | Duration of vigorous activity | 914 | duration_vigorous_activity | Collapsed at 3.5 SD above mean, log-transformed |
| Smoking | Smoking status | 20116 | smoking_status | Binarized into yes and no |
| | Past tobacco smoking | 1249 | past_tobacco | Reverse coding |
| | Pack years | 20161 | pack_years | Never smokers set to zero using data field 20160, log-transformed |

*Note.* Table shows the categories of the variables, the name and field ID of the variables in the UK Biobank data, the variable name as used in the present study and as seen in figure 4, and transformations that were performed to obtain the final variables.
